# Diagnostic Performance Comparison between Generative AI and Physicians: A Systematic Review and Meta-Analysis

**DOI:** 10.1101/2024.01.20.24301563

**Authors:** Hirotaka Takita, Daijiro Kabata, Shannon L Walston, Hiroyuki Tatekawa, Kenichi Saito, Yasushi Tsujimoto, Yukio Miki, Daiju Ueda

## Abstract

**Background:** The rapid advancement of generative artificial intelligence (AI) has led to the wide dissemination of models with exceptional understanding and generation of human language. Their integration into healthcare has shown potential for improving medical diagnostics, yet a comprehensive diagnostic performance evaluation of generative AI models and the comparison of their diagnostic performance with that of physicians has not been extensively explored.

**Methods:** In this systematic review and meta-analysis, a comprehensive search of Medline, Scopus, Web of Science, Cochrane Central, and MedRxiv was conducted for studies published from June 2018 through December 2023, focusing on those that validate generative AI models for diagnostic tasks. The risk of bias was assessed using the Prediction Model Study Risk of Bias Assessment Tool. Meta-regression was performed to summarize the performance of the models and to compare the accuracy of the models with that of physicians.

**Results:** The search resulted in 54 studies being included in the meta-analysis. Nine generative AI models were evaluated across 17 medical specialties. The quality assessment indicated a high risk of bias in the majority of studies, primarily due to small sample sizes. The overall accuracy for generative AI models across 54 studies was 56.9% (95% confidence interval [CI]: 51.0–62.7%). The meta-analysis demonstrated that, on average, physicians exceeded the accuracy of the models (difference in accuracy: 14.4% [95% CI: 4.9–23.8%], p-value =0.004). However, both Prometheus (Bing) and GPT-4 showed slightly better performance compared to non-experts (-2.3% [95% CI: -27.0–22.4%], p-value = 0.848 and -0.32% [95% CI: -14.4–13.7%], p-value = 0.962), but slightly underperformed when compared to experts (10.9% [95% CI: -13.1–35.0%], p-value = 0.356 and 12.9% [95% CI: 0.15–25.7%], p-value = 0.048). The sub-analysis revealed significantly improved accuracy in the fields of Gynecology, Pediatrics, Orthopedic surgery, Plastic surgery, and Otolaryngology, while showing reduced accuracy for Neurology, Psychiatry, Rheumatology, and Endocrinology compared to that of General Medicine. No significant heterogeneity was observed based on the risk of bias.

**Conclusions:** Generative AI exhibits promising diagnostic capabilities, with accuracy varying significantly by model and medical specialty. Although they have not reached the reliability of expert physicians, the findings suggest that generative AI models have the potential to enhance healthcare delivery and medical education, provided they are integrated with caution and their limitations are well-understood.

**Key Points:** **Question:** What is the diagnostic accuracy of generative AI models and how does this accuracy compare to that of physicians?

**Findings:** This meta-analysis found that generative AI models have a pooled accuracy of 56.9% (95% confidence interval: 51.0–62.7%). The accuracy of expert physicians exceeds that of AI in all specialties, however, some generative AI models are comparable to non-expert physicians.

**Meaning:** The diagnostic performance of generative AI models suggests that they do not match the level of experienced physicians but that they may have potential applications in healthcare delivery and medical education.

## Introduction

In recent years, the advent of generative artificial intelligence (AI) has marked a transformative era in our society.^1–8^ These advanced computational systems have demonstrated exceptional proficiency in interpreting and generating human language, thereby setting new benchmarks in AI’s capabilities. Generative AI, with their deep learning architectures, have rapidly evolved, showcasing a remarkable understanding of complex language structures, contexts, and even images. This evolution has not only expanded the horizons of AI but also opened new possibilities in various fields, including healthcare.^9^

The integration of generative AI models in the medical domain has spurred a growing body of research focusing on their diagnostic capabilities.^10^ Studies have extensively examined the performance of these models in interpreting clinical data, understanding patient histories, and even suggesting possible diagnoses.^11,12^ In medical diagnosis, the accuracy, speed, and efficiency of generative AI models in processing vast amounts of medical literature and patient information have been highlighted, positioning them as valuable tools. This research has begun to outline the strengths and limitations of generative AI models in diagnostic tasks in healthcare.

Despite the growing research on generative AI models in medical diagnostics, there remains a significant gap in the literature: a comprehensive meta-analysis of the diagnostic capabilities of the models, followed by a comparison of their performance with that of physicians. Such a comparison is crucial for understanding the practical implications and effectiveness of generative AI models in real-world medical settings. While individual studies have provided insights into the capabilities of generative AI models,^13,14^ a systematic review and meta-analysis is necessary to aggregate these findings and draw more robust conclusions about their comparative effectiveness against traditional diagnostic practices by physicians.

This paper aims to bridge the existing gap in the literature by conducting a meticulous meta-analysis of the diagnostic capabilities of generative AI models in healthcare. Our focus is to provide a comprehensive diagnostic performance evaluation of generative AI models and compare their diagnostic performance with that of physicians. By synthesizing the findings from various studies, we endeavor to offer a nuanced understanding of the effectiveness, potential, and limitations of generative AI models in medical diagnostics. This analysis is intended to serve as a foundational reference for future research and practical applications in the field, ultimately contributing to the advancement of AI-assisted diagnostics in healthcare.

## Methods

### Protocol and Registration

This systematic review was prospectively registered with PROSPERO (CRD42023494733). Our study adhered to the relevant sections of guidelines from the Preferred Reporting Items for a Systematic Review and Meta-analysis (PRISMA) of Diagnostic Test Accuracy Studies.^15,16^ All stages of the review (title and abstract screening, full-text screening, data extraction, and assessment of bias) were performed in duplicate by two independent reviewers (H.Takita and D.U.), and disagreements were resolved by discussion with a third independent reviewer (H.Tatekawa).

### Search Strategy and Study Selection

A search was performed to identify studies that validate a generative AI model for diagnostic tasks. A search strategy was developed, including variations of the terms *generative AI* and *diagnosis*. The search strategy was as follows: articles in English that included the words “large language model”, “LLM”, “generative artificial intelligence”, “generative AI”, “generative pre-trained transformers”^1^, “GPT”, “Bing”, “Prometheus”, “Bard”, “PaLM”^6,7^, “Pathways Language Model”, “LaMDA”^8^, “Language Model for Dialogue Applications”, “Llama”^4,5^, or “Large Language Model Meta AI” and also “diagnosis”, “diagnostic”, “quiz”, “examination”, or “vignette” were included. We searched the following electronic databases for literature from June 2018 through December 2023: Medline, Scopus, Web of Science, Cochrane Central, and MedRxiv. June 2018 represents when the first generative AI model was published.^1^ We included all articles that fulfilled the following inclusion criteria: primary research studies that validate a generative AI for diagnosis. We applied the following exclusion criteria to our search: review articles, case reports, comments, editorials, and retracted articles.

### Data Extraction

Titles and abstracts were screened before full-text screening. Data was extracted using a predefined data extraction sheet. A count of excluded studies, including the reason for exclusion, was recorded in a PRISMA flow diagram.^16^ We extracted information from each study including the first author, model with its version, model task, test dataset type (internal, external, or unknown),^17^ medical specialty, accuracy, sample size, and publication status (pre-print or peer-reviewed) for the meta-analysis of generative AI performance. Most generative AI models only presented their training period without any information on which data were used for training. Therefore, when generative AI models were tested with data outside of the training period, the test dataset type was classified as external testing, and when tested with data that were publicly available during the training period, it was classified as unknown. In addition to this, when both the model and the physician’s diagnostic performance were presented in the same paper, we extracted both for meta-analysis. We also considered the type of physician involved in relevant studies. We classified physicians as non-experts if they were trainees or residents. In contrast, those beyond this stage in their career were categorized as experts. When a single model used multiple prompts and individual performances were available in one article, we took the average of them.

### Quality Assessment

We used the Prediction Model Study Risk of Bias Assessment Tool (PROBAST) to assess papers for bias and applicability.^17^ This tool uses signaling questions in four domains (participants, predictors, outcomes, and analysis) to provide both an overall and a granular assessment. We did not include some PROBAST signaling questions because they are not relevant to generative AI models. Details of modifications made to PROBAST are in Appendix Table S1 (online).

### Statistical Analysis

We calculated the pooled accuracy of diagnosis brought by generative AI models and physicians based on the previously reported studies. The pooled diagnosis accuracies were compared between all AI models and overall physicians using the multivariable random-effect meta-regression model with adjustment for medical speciality, task of models, type of test dataset, level of bias, and publication status. In addition to the comparison of all AI models and overall physicians, we compared each AI model with overall physicians and each AI model with each physician experience level (expert or non-expert). Furthermore, we assessed the variation of generative AI model accuracy across specialities. For fitting the meta-regression models, a restricted maximum likelihood estimator was utilized with the “metafor” package in R. To assess the impact of publication bias on the comparison of the diagnosis performance between the AI models and the physicians, we used a funnel plot and Egger’s regression test. All statistical analyses were conducted using R version 4.3.0.

## Results

### Study Selection and Characteristics

We identified 13,966 studies, of which 7,940 were duplicates. After screening, 54 studies were included in the meta-analysis^11–14,19–68^ (Figure 1 and Table 1). The most evaluated models were GPT-4^3^ (31 articles) and GPT-3.5^2^ (28), while models such as GPT-4V^69^ (6), PaLM2^7^ (3), Llama 2^5^ (2), Prometheus (2), GPT-3^2^ (1), Glass AI^70^ (1), and Med-42^56^ (1) had less representation. GPT-3, GPT-3.5, GPT-4, and GPT-4V are available in ChatGPT or its application programming interface (Open AI, San Francisco, CA). PaLM2 is implemented in Bard (Google, Menlo Park, CA). Bing (Microsoft, Redmond, WA) incorporates Prometheus, which is based on OpenAI’s GPT technology. Med-42 is a fine-tuned version of the open-source large language model, Llama 2 (Meta, Menlo Park, CA). Lastly, Glass AI is implemented in Glass (Glass Health, San Francisco, CA). The review spanned a wide range of medical specialties, with General medicine being the most common (14 articles). Other specialties like Radiology (10), Ophthalmology (8), Emergency medicine (5), Neurology (3), and Dermatology (3) were represented, as well as Gastroenterology, Cardiology, Pediatrics, Otolaryngology, Urology, Endocrinology, Gynecology, Orthopedic surgery, Rheumatology, Psychiatry, and Plastic surgery with one article each. Regarding model tasks, free text tasks were the most common, with 47 articles, followed by choice tasks at 13. For test dataset types, 40 articles involved external testing, while 14 were unknown because the training data for the generative AI models was unknown. Of the included studies, 37 were peer-reviewed, while 17 were preprints. Study characteristics are shown in Table 1 and Appendix Table S2 (online). Thirteen studies compared the performance of generative AI models with that of physicians.^30,31,33–39,47,50,54,58^ GPT-4 (8 articles) was the most frequently compared with physicians, followed by GPT-3.5 (7), GPT-4V (2), Llama 2 (1), and GPT-3 (1). While comparisons between both expert and non-expert physicians were found for GPT-4, GPT-3.5, GPT-4V, and GPT-3, only comparisons with experts were found for Llama 2, with no comparisons involving non-experts.

**Figure 1:**
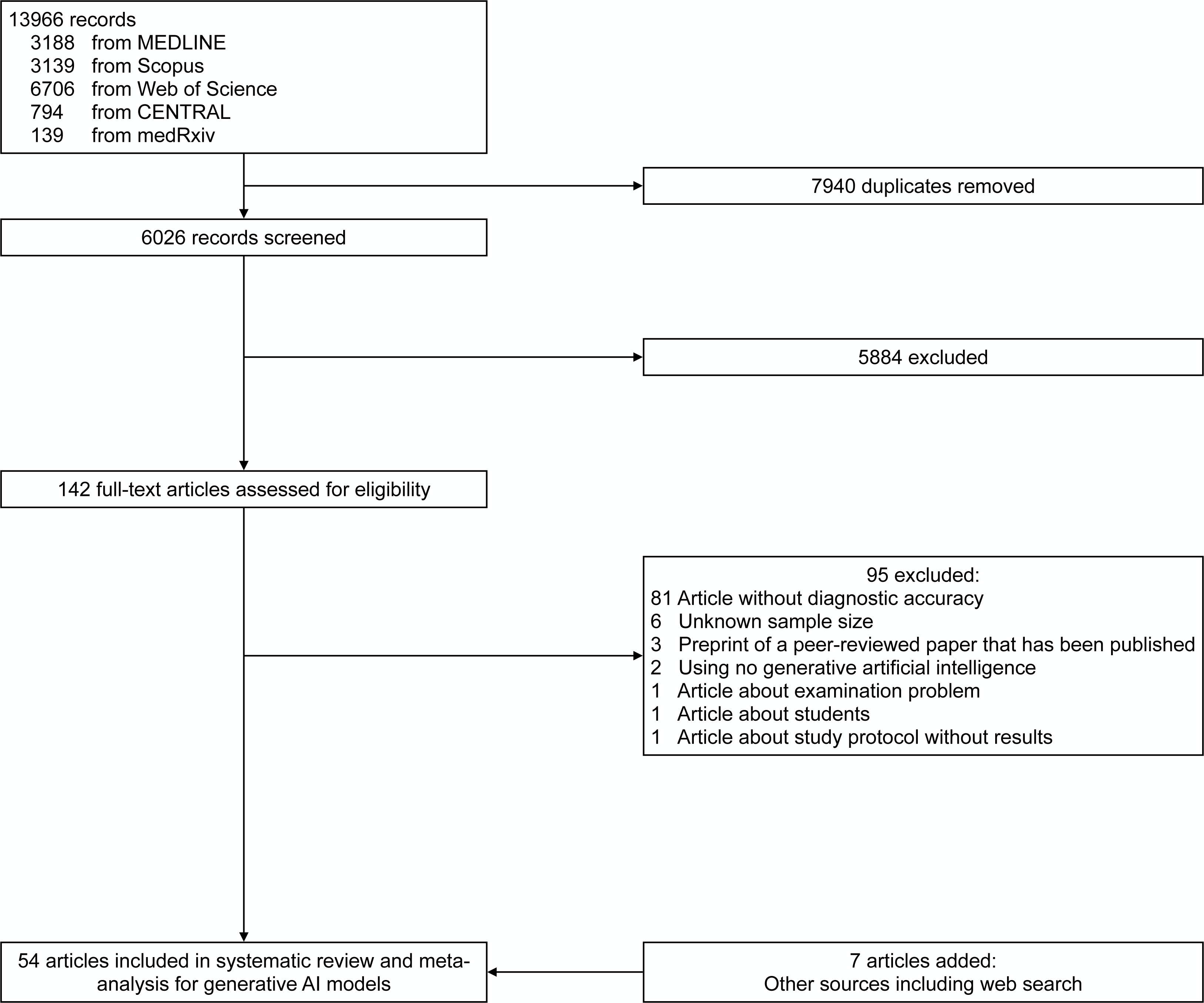
Eligibility criteria.

**Table 1:** Study characteristics.

| Citation | First author | Model | Model task | Test type | Specialty | Comparison group | Cases | Publication status | Overall risk of bias | Overall applicability |
| --- | --- | --- | --- | --- | --- | --- | --- | --- | --- | --- |
| <sup>11</sup> | Ueda | GPT-4 | Free text | External | Radiology | NA | 313 | Peer-reviewed | Low | High |
| <sup>12</sup> | Kanjee | GPT-4 | Free text | External | General medicine | NA | 70 | Peer-reviewed | High | Low |
| <sup>13</sup> | Hirosawa | PaLM2 | Free text | External | General medicine | NA | 82 | Peer-reviewed | High | Low |
| <sup>14</sup> | Shea | GPT-4 | Free text | External | General medicine | NA | 6 | Peer-reviewed | High | Low |
| <sup>19</sup> | Chee | GPT-3.5 | Free text | External | Otolaryngology | NA | 7 | Peer-reviewed | High | Low |
| <sup>20</sup> | Lyons | Prometheus, GPT-4 | Free text, Choice | External | Ophthalmology | NA | 44 | Peer-reviewed | High | Low |
| <sup>21</sup> | Hirosawa | GPT-3.5, GPT-4 | Free text | Unknown | General medicine | NA | 52 | Peer-reviewed | High | Low |
| <sup>22</sup> | Benoit | GPT-3.5 | Free text, Choice | Unknown | General medicine | NA | 45 | Preprint | High | Low |
| <sup>23</sup> | Hirosawa | GPT-3.5 | Free text | External | General medicine | NA | 30 | Peer-reviewed | High | Low |
| <sup>24</sup> | Wei | GPT-4 | Choice | External | Psychiatry | NA | 60 | Peer-reviewed | High | Low |
| <sup>25</sup> | Ueda | GPT-4 | Free text | External | General medicine | NA | 62 | Preprint | High | High |
| <sup>26</sup> | Allahqoli | GPT-3.5 | Free text | Unknown | Gynecology | NA | 30 | Peer-reviewed | High | Low |
| <sup>27</sup> | Levartovsky | GPT-4 | Choice | External | Gastroenterology | NA | 20 | Peer-reviewed | High | Low |
| <sup>28</sup> | Bushuven | GPT-3.5, GPT-4 | Free text, Choice | External | Emergency medicine | NA | 22 | Peer-reviewed | High | Low |
| <sup>29</sup> | Knebel | GPT-3.5 | Free text, Choice | External | Ophthalmology | NA | 10 | Peer-reviewed | High | Low |
| <sup>30</sup> | Mitsuyama | GPT-4 | Free text | External | Radiology | Expert, Non-expert | 99 | Preprint | High | Low |
| <sup>31</sup> | Pillai | GPT-3.5, GPT-4, Llama 2 | Free text | Unknown | Endocrinology | Expert | 20 | Peer-reviewed | High | Low |
| <sup>32</sup> | Brin | GPT-4V | Free text | External | Radiology | NA | 36 | Preprint | High | Low |
| <sup>33</sup> | Horiuchi | GPT-4 | Free text | External | Radiology | Expert, Non-expert | 30 | Preprint | High | High |
| 34 | Ito | GPT-4 | Free text,<br>Choice | Unknown | General medicine | Expert | 45 | Peer-reviewed | High | Low |
| 35 | Horiuchi | GPT-4,<br>GPT-4V | Free text | External | Radiology | Expert,<br>Non-expert | 106 | Preprint | Low | High |
| 36 | Madadi | GPT-3.5,<br>GPT-4 | Free text | Unknown | Ophthalmology | Expert | 22 | Preprint | High | Low |
| 37 | Sorin | GPT-4V | Free text | External | Ophthalmology | Non-expert | 40 | Preprint | High | Low |
| 38 | Delsoz | GPT-3.5,<br>GPT-4 | Free text | Unknown | Ophthalmology | Expert | 20 | Preprint | High | Low |
| 39 | Levine | GPT-3 | Free text,<br>Choice | External | General medicine | Expert | 48 | Preprint | High | Low |
| 40 | Schubert | GPT-4V | Free text | External | General medicine | NA | 93 | Preprint | High | High |
| 41 | Sultan | GPT-3.5 | Free text | External | Pediatrics | NA | 30 | Peer-reviewed | High | Low |
| 42 | Kiyohara | PaLM2,<br>GPT-3.5,<br>GPT-4 | Choice | Unknown | Cardiology | NA | 66 | Preprint | High | Low |
| 43 | Horiuchi | GPT-4 | Free text | External | Neurology | NA | 100 | Peer-reviewed | Low | High |
| 44 | Stoneham | GPT-4 | Free text | External | Dermatology | NA | 36 | Peer-reviewed | High | Low |
| 45 | Rundle | GPT-3.5 | Free text | External | Dermatology | NA | 39 | Peer-reviewed | High | Low |
| 46 | Rojas-Carabali | GPT-3.5,<br>GPT-4,<br>Glass AI | Free text | External | Ophthalmology | NA | 6 | Peer-reviewed | High | Low |
| 47 | Fraser | GPT-3.5,<br>GPT-4 | Free text | Unknown | Emergency<br>medicine | Expert | 30 | Peer-reviewed | High | Low |
| 48 | Krusche | GPT-4 | Free text | External | Rheumatology | NA | 132 | Peer-reviewed | Low | Low |
| 49 | Galetta | GPT-4 | Free text | External | Neurology | NA | 24 | Peer-reviewed | High | Low |
| 50 | Delsoz | GPT-3.5 | Free text | Unknown | Ophthalmology | Non-expert | 11 | Peer-reviewed | High | Low |
| 51 | Hu | GPT-4 | Free text | Unknown | Ophthalmology | NA | 10 | Peer-reviewed | High | Low |
| 52 | Abi-Rafeh | GPT-3.5 | Free text | External | Plastic surgery | NA | 16 | Peer-reviewed | High | Low |
| 53 | Koga | PaLM2,<br>GPT-3.5,<br>GPT-4 | Free text | External | Neurology | NA | 25 | Peer-reviewed | High | Low |
| 54 | Xv | GPT-3.5 | Free text | External | Urology | Non-expert | 306 | Peer-reviewed | Low | Low |
| 55 | Reese | GPT-4 | Free text | External | General medicine | NA | 75 | Preprint | High | Low |
| 56 | Han | GPT-3.5,<br>GPT-4,<br>GPT-4V,<br>Llama 2,<br>Med-42 | Choice | Unknown | General medicine | NA | 140,<br>348 | Preprint | Unclear | High |
| 57 | Senkaiahliyan | GPT-4V | Free text | Unknown | Radiology | NA | 69 | Preprint | High | Low |
| 58 | Williams | GPT-3.5 | Choice | External | Emergency<br>medicine | Non-expert | 500 | Preprint | Low | Low |
| 59 | Tenner | GPT-3.5 | Free text | External | Radiology | NA | 40 | Preprint | High | Low |
| 60 | Mori | GPT-4 | Choice | External | Radiology | NA | 151 | Peer-reviewed | Low | Low |
| 61 | Mykhalko | GPT-3.5 | Free text | External | General medicine | NA | 50 | Peer-reviewed | High | High |
| 62 | Andrade-Castellanos | GPT-3.5 | Free text | External | General medicine | NA | 10 | Peer-reviewed | High | High |
| 63 | Daher | GPT-3.5 | Free text | External | Orthopedic<br>surgery | NA | 29 | Peer-reviewed | High | Low |
| 64 | Suthar | GPT-4 | Free text | External | Radiology | NA | 140 | Peer-reviewed | Low | High |
| 65 | Nakaura | Prometheus,<br>GPT-3.5 | Free text | External | Radiology | NA | 28 | Peer-reviewed | High | Low |
| 66 | Berg | GPT-3.5,<br>GPT-4 | Free text | External | Emergency<br>medicine | NA | 30 | Peer-reviewed | High | Low |
| 67 | Gebrael | GPT-4 | Choice | External | Emergency<br>medicine | NA | 56 | Peer-reviewed | High | Low |
| 68 | Ravipati | GPT-3.5 | Free text | Unknown | Dermatology | NA | 32 | Peer-reviewed | High | Low |

### Quality Assessment

PROBAST assessment led to an overall rating of 45/54 (83%) studies at high risk of bias, 8/54 (15%) studies at low risk of bias, 10/54 (19%) studies at high concern for generalizability, and 44/54 (81%) studies at low concern for generalizability (Figure 2). The main factors of this evaluation were studies that evaluated models with a small test set and studies that cannot prove external evaluation due to the unknown training data of generative AI models. Detailed results are shown in Appendix Table S2 (online).

**Figure 2:**
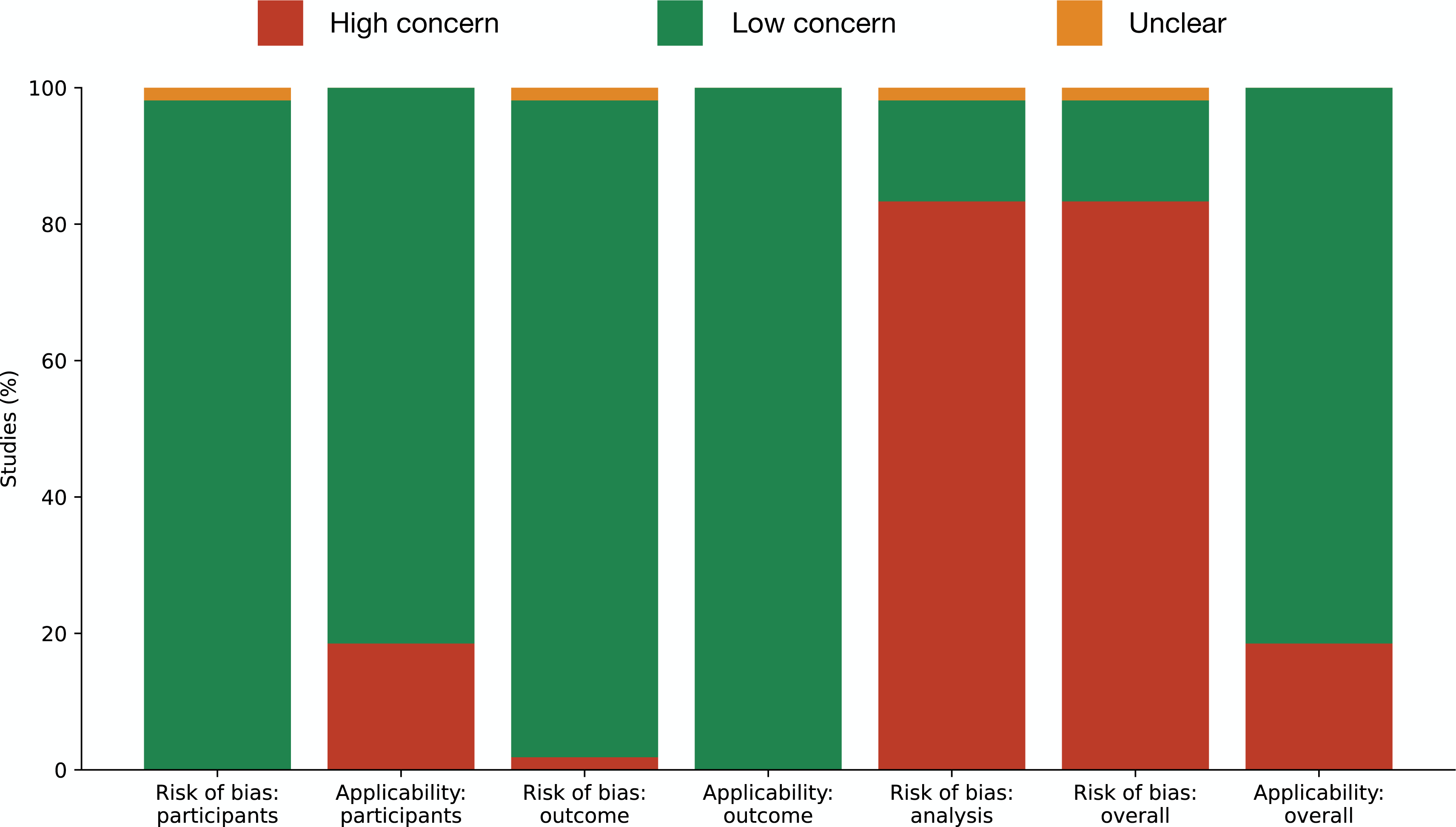
Summary of Prediction Model Study Risk of Bias Assessment Tool (PROBAST) risk of bias. Assessment of risk of biases using the PROBAST tool for generative AI model studies included in the meta-analysis (N = 54). The participants and the outcome determination were predominantly at low risk of bias, but there was a high risk of bias for analysis (83%) and the overall evaluation (83%). Applicability for participants and outcomes shows a predominantly low concern, whereas overall applicability has 19% high concern.

### Meta-analysis

The overall accuracy for generative AI models was found to be 56.9% with a 95% CI of 51.0–62.7%. In the meta-regression, we observed that physicians generally outperformed generative AI models in various scenarios (Figure 3). This superiority was evident when comparing AI models to overall physician performance, where physicians demonstrated a significant 14.4% higher performance on average (95% CI: 4.9–23.8%, p =0.004). Interestingly, when comparing the performance of the Prometheus and GPT-4 models against non-experts, both models demonstrated a slight but not statistically significant superiority, with differences of -2% (95% CI: -27.0 to 22.4%, p = 0.848) and -0.3% (95% CI: -14.4 to 13.7%, p = 0.962), respectively. However, both models underperformed in comparison to experts, showing a 10.9% difference [95% CI: -13.1 to 35.0%, p-value = 0.356 for Prometheus] and 12.9% [95% CI: 0.15 to 25.6%, p-value = 0.048 for GPT-4]. The performance of all models but Prometheus and GPT-4 was inferior to both experts and non-experts in all comparisons. GPT-3, GPT-3.5, and PaLM2 were significantly inferior only when compared to expert physicians, whereas Llama 2, Glass, and Med-42 demonstrated substantial inferiority against both expert and non-expert physicians (p-values < 0.05).

**Figure 3:**
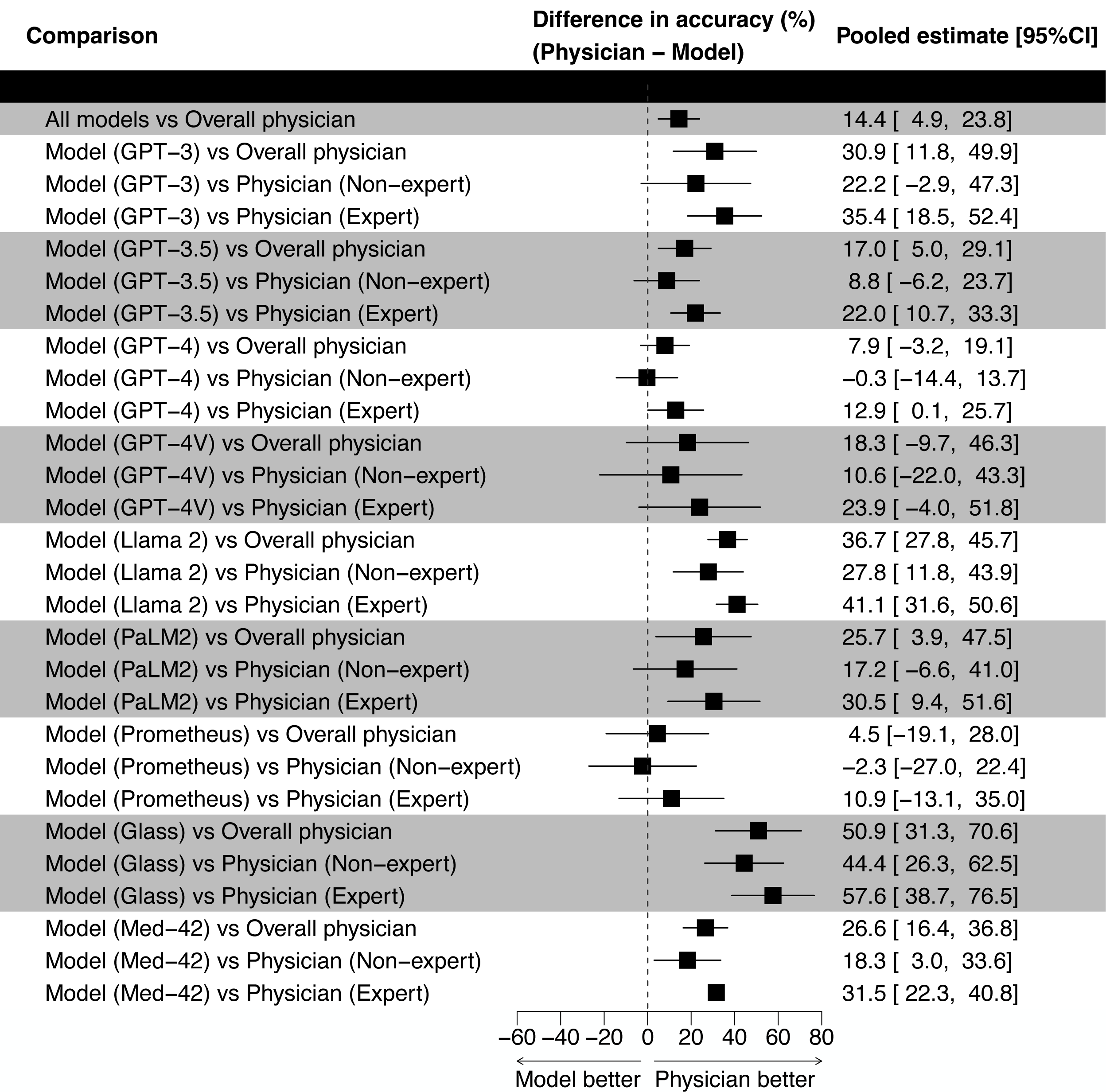
Comparison results between models and physicians. This figure demonstrates the differences in accuracy between various AI models and physicians. It specifically compares the performance of AI models against the overall accuracy of physicians, as well as against non-experts and experts separately. Each horizontal line represents the range of accuracy differences for the model compared to the physician category. The percentage values displayed on the right-hand side correspond to these mean differences, with the values in parentheses providing the 95% confidence intervals for these estimates. The dotted vertical line marks the 0% difference threshold, indicating where the model’s accuracy is exactly the same as that of the physicians’. Positive values (to the right of the dotted line) suggest that the physicians outperformed the model, whereas negative values (to the left) indicate that the model was more accurate than the physicians.

**Figure 4:**
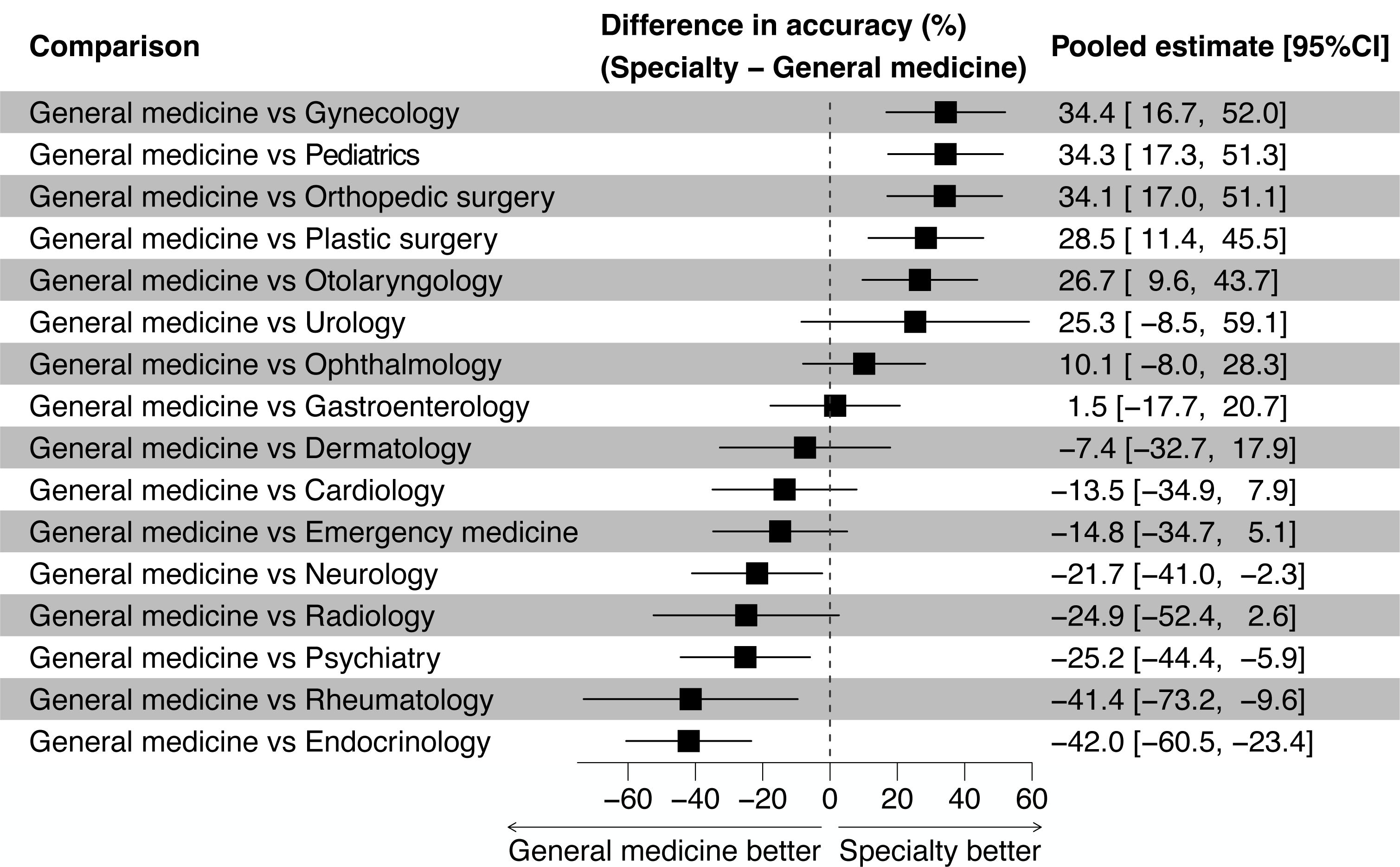
Generative AI performance among specialities. This figure demonstrates the differences in accuracy of generative AI models for specialties. Each horizontal line represents the range of accuracy differences between the speciality and General medicine. The percentage values displayed on the right-hand side correspond to these mean differences, with the values in parentheses providing the 95% confidence intervals for these estimates. The dotted vertical line marks the 0% difference threshold, indicating where the performance of generative AI models in the speciality is exactly the same as that of General medicine. Positive values (to the right of the dotted line) suggest that the model performance for the speciality was greater than that for General medicine, whereas negative values (to the left) indicate that the model performance for the speciality was less than that for General medicine.

In our meta-regression, we also found a remarkable difference in accuracy, with significant improvements in several specialties compared with General medicine. Specifically, AI performance in Gynecology, Pediatrics, Orthopedic surgery, Plastic surgery, and Otolaryngology outpace General medicine significantly, exhibiting differences of 34.4% (95% CI: 16.7–52.0%, p < 0.001), 34.3% (95% CI: 17.3–51.4%, p < 0.001), 34.1% (95% CI: 17.0–51.1%, p < 0.001), 28.5% (95% CI: 11.4–45.5%, p = 0.002), and 26.7% (95% CI: 9.6–43.7%, p = 0.004) respectively. Conversely, General medicine outperformed some areas such as Neurology, Psychiatry, Rheumatology, and Endocrinology. These areas witnessed a decline in accuracy with differences of -21.7% (95% CI: -41.0 to -2.3%, p = 0.030) in Neurology, -25.1% (95% CI: -44.4 to -5.9%, p = 0.012) in Psychiatry, -41.4% (95% CI: -73.2 to -9.6%, p = 0.013) in Rheumatology, and the most notable decrease in Endocrinology with -42.0% (95% CI: -60.5 to -23.4%, p < 0.001). These findings suggest that generative AI’s performance is not uniform across all medical specialties, highlighting the necessity for specialty-specific optimization to harness its full potential effectively. No significant difference was observed based on the risk of bias (p = 0.77) or based on publication status (p = 0.58). We assessed publication bias by using a regression analysis to quantify funnel plot asymmetry (Appendix Figures S1 [online]), and it suggested a risk of publication bias (p = 0.027).

## Discussion

In this systematic review and meta-analysis, we analyzed the diagnostic performance of generative AI and physicians. We initially identified 13,966 studies, ultimately including 54 in the meta-analysis. The study spanned various AI models and medical specialties, with GPT-4 being the most evaluated. Quality assessment revealed a majority of studies at high risk of bias. The meta-analysis showed a pooled accuracy of 57% (95% CI: 51–63%) for generative AI models. Physicians generally outperformed AI models, although in non-expert settings, some AI models showed comparable performance. Our analysis also highlighted significant differences in effectiveness across medical fields. To the best of our knowledge, this is the first meta-analysis of generative AI models in diagnostic tasks. This comprehensive study highlights the varied capabilities and limitations of generative AI in medical diagnostics.

The meta-analysis of generative AI models in healthcare reveals crucial insights for clinical practice. Despite the overall modest accuracy of 57% for generative AI models in medical applications, this suggests its potential utility in certain clinical scenarios. The variation in effectiveness across specialties, particularly the lower effectiveness in some fields underscores the need for cautious implementation and further refinement of AI models in these areas. The data indicates that generative AI models possess a propensity towards knowledge in some medical specialties, and by understanding and utilizing their characteristics, they have the potential to function as a valuable support tool in medical settings. Importantly, the similar performance of Prometheus and GPT-4 to physicians in non-expert scenarios highlights the possibility of AI augmenting healthcare delivery in resource-limited settings or as a preliminary diagnostic tool, thereby potentially increasing accessibility and efficiency in patient care.^71^

The studies comparing generative AI and physician performance, particularly in the context of medical education, offer intriguing perspectives.^72^ The overall higher accuracy of physicians compared to AI models emphasizes the irreplaceable value of human judgement and experience in medical decision-making. However, the comparable performance of Prometheus and GPT-4 to physicians in non-expert settings reveals an opportunity for integrating AI into medical training. This could include using AI as a teaching aid for medical students and residents, especially in simulating non-expert scenarios where AI’s performance is nearly equivalent to that of healthcare professionals.^73^ Such integration could enhance learning experiences, offering diverse clinical case studies and facilitating self-assessment and feedback. Additionally, the narrower performance gap between some generative AI models and physicians even in expert settings suggests that AI could be used to supplement advanced medical education, helping to identify areas for improvement and providing supporting information. This approach could foster a more dynamic and adaptive learning environment, preparing future medical professionals for an increasingly digital healthcare landscape.

Although there are no statistically significant differences in diagnostic performance among the risks of bias, the PROBAST quality assessment reveals a high risk of bias in 83% of studies.^17^ This raises significant concerns about the reliability of current generative AI research in healthcare. This highlights the crucial need for rigorous and transparent methodologies, including the necessity of large amounts of external evaluation to assess real-world performance accurately.^74^ Moreover, the transparency of training data and its collection period is paramount. Without this transparency, it is impossible to determine whether the test dataset is an external dataset or not. Transparency ensures an understanding of the model’s knowledge, context, and limitations, aids in identifying potential biases, and facilitates independent replication and validation, which are fundamental to scientific integrity. As generative AI continues to evolve, fostering a culture of rigorous transparency is essential to ensure their safe, effective, and equitable application in clinical settings,^75^ ultimately enhancing the quality of healthcare delivery and medical education.

The methodology of this study, while comprehensive, has limitations. The performance of generative AI models might vary significantly in real-world scenarios, which are often more complex than research settings. There were not many studies that compared generative AI and physicians using the same sample. Future research should focus on addressing limitations by conducting studies with more diverse datasets, exploring the performance of generative AI models in varied clinical environments, and examining their impact on different patient demographics. Additionally, investigating the intersecting impact of physicians using generative AI models clinically, such as changes in performance, would be valuable.

In conclusion, this meta-analysis provides a nuanced understanding of the capabilities and limitations of generative AI in medical diagnostics. While generative AI models, particularly advanced iterations like Prometheus and GPT-4, have shown progressive improvements and hold promise for assisting in diagnosis, their effectiveness remains highly variable across different models and medical specialties. With an overall moderate accuracy of 57%, generative AI models are not yet reliable substitutes for expert physicians but may serve as valuable aids in non-expert scenarios and as educational tools for medical trainees. The findings also underscore the need for continued advancements and specialization in model development, as well as rigorous, externally validated research to overcome the prevalent high risk of bias and ensure generative AIs’ effective integration into clinical practice. As the field evolves, continuous learning and adaptation for both generative AI models and medical professionals are imperative, alongside a commitment to transparency and stringent research standards. This approach will be crucial in harnessing the potential of generative AI models to enhance healthcare delivery and medical education while safeguarding against their limitations and biases.

## Data Availability

All data produced in the present study are available upon reasonable request to the authors.

## Acknowledgement

We utilized ChatGPT for assistance with parts of the English proofing.

## IRB Approval

Not applicable.

## Reproducible Research Statement

Study protocol and metadata are available from Dr. Ueda (e-mail).

