## Appendix for "Diagnostic Performance Comparison between Generative AI and Physicians: A Systematic Review and Meta-Analysis"

**Table of Contents:**

**Section S1: Supplementary Figures**

Appendix Figure S1: Funnel plot

**Section S2: Supplementary Tables**

Appendix Table S1: PROBAST modifications

Appendix Table S2: Detailed study characteristics

**Section S1: Supplementary Figures**

Appendix Figure S1: Funnel plot

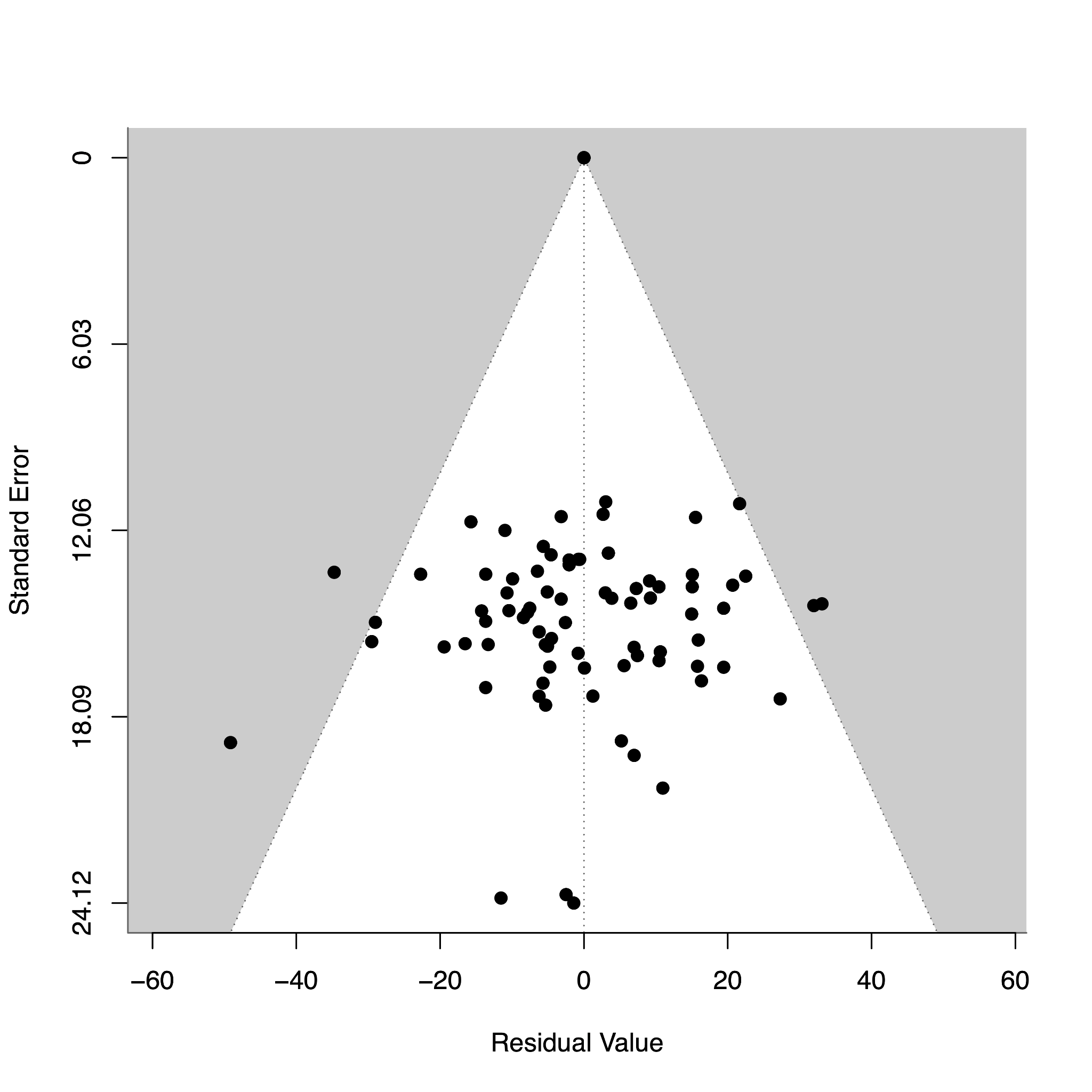

The funnel plot illustrates the distribution of the residuals of the fitted values corresponding to their standard errors in the meta-regression. The Egger test results z-value = -2.2094 and p = 0.0271, which indicates the possible presence of publication bias.

**Section S2: Supplementary Tables**

Appendix Table S1: PROBAST modifications

| PROBAST Items | Modifications |
| --- | --- |
| Domain 1: Participants | No changes (Refer to participant data for diagnosis) |
| Domain 2: Predictors | N/A–removed from scoring |
| Domain 3: Outcome | Items 3.3, 3.5, and 3.6 N/A |
| Domain 4: Analysis | Items 4.5, 4.6, and 4.9 N/A |
| Domain 5: Overall | No changes |

Appendix Table S2: Detailed study characteristics

| **Citation** | **Publication year** | **First author** | **Model** | **Version** | **Database explanation** | **Reference standard** | **ROB for participants** | **Applicability for participants** | **ROB for outcome** | **Applicability for outcome** | **ROB for analysis** |
| --- | --- | --- | --- | --- | --- | --- | --- | --- | --- | --- | --- |
| ^11^ | 2023 | Ueda | GPT-4 | Not written | The quiz “Diagnosis Please” in Radiology | Answer | Low | High | Low | Low | Low |
| ^12^ | 2023 | Kanjee | GPT-4 | Not written | Clinical vignettes from case reports in NEJM | Expert consensus | Low | Low | Low | Low | High |
| ^13^ | 2023 | Hirosawa | PaLM2 | Not written | Clinical vignettes representing various common complaints | Expert consensus | Low | Low | Low | Low | High |
| ^14^ | 2023 | Shea | GPT-4 | Not written | Clinical vignettes about patients who had delay of definitive diagnosis longer than 1 month | Expert consensus | Low | Low | Low | Low | High |
| ^19^ | 2023 | Chee | GPT-3.5 | Not written | Hypothetical clinical vignettes representing vertigo | Expert consensus | Unclear | Low | High | Low | High |
| ^20^ | 2023 | Lyons | Prometheus, GPT-4 | Not written | Hypothetical clinical vignettes representing common ophthalmic complaints | Expert consensus | Low | Low | Low | Low | High |
| ^21^ | 2023 | Hirosawa | GPT-3.5, GPT-4 | March 14, 2023, March 25, 2023 | Clinical vignettes about general internal medicine | Expert consensus | Low | Low | Low | Low | High |
| ^22^ | 2023 | Benoit | GPT-3.5 | January 9, 2023 | Clinical vignettes representing various common complaints | Expert consensus | Low | Low | Unclear | Low | High |
| ^23^ | 2023 | Hirosawa | GPT-3.5 | December 15, 2023 | Clinical vignettes about general internal medicine | Expert consensus | Low | Low | Low | Low | High |
| ^24^ | 2023 | Wei | GPT-4 | Not written | Clinical vignettes about pediatrics | Expert consensus | Low | Low | Low | Low | High |
| ^25^ | 2023 | Ueda | GPT-4 | March 23, 2023 | The quiz “Image Challenge” in NEJM | Answer | Low | High | Low | Low | High |
| ^26^ | 2023 | Allahqoli | GPT-3.5 | Not written | Clinical vignettes about obstetrics and gynecology | Expert consensus | Low | Low | Low | Low | High |
| ^27^ | 2023 | Levartovsky | GPT-4 | Not written | Clinical vignettes about patients with ulcerative colitis | Expert consensus | Low | Low | Low | Low | High |
| ^28^ | 2023 | Bushuven | GPT-3.5, GPT-4 | March 23, 2023 | Clinical vignettes about emergency medicine | Expert consensus | Low | Low | Low | Low | High |
| ^29^ | 2023 | Knebel | GPT-3.5 | March 14, 2023 | Hypothetical clinical vignettes representing acute ocular symptoms | Expert consensus | Low | Low | Low | Low | High |
| ^30^ | 2023 | Mitsuyama | GPT-4 | May 24, 2023 | Clinical vignettes about imaging findings of brain tumors | Expert consensus | Low | Low | Low | Low | High |
| ^31^ | 2023 | Pillai | GPT-3.5, GPT-4, Llama 2 | Not written | Clinical vignettes about FMF and DIRA | Expert consensus | Low | Low | Low | Low | High |
| ^32^ | 2023 | Brin | GPT-4V | Not written | Clinical vignettes about imaging findings | Expert consensus | Low | Low | Low | Low | High |
| ^33^ | 2023 | Horiuchi | GPT-4 | May 24, 2023 | The quiz "Freiburg Neuropathology Case Conference" in Clinical Neuroradiology | Answer | Low | High | Low | Low | High |
| ^34^ | 2023 | Ito | GPT-4 | March 14, 2023 | Clinical vignettes representing various common complaints | Expert consensus | Low | Low | Low | Low | High |
| ^35^ | 2023 | Horiuchi | GPT-4, GPT-4V | September 25, 2023 | The quiz "Test yourself" in Skeletal Radiology | Answer | Low | High | Low | Low | Low |
| ^36^ | 2023 | Madadi | GPT-3.5, GPT-4 | Not written | Clinical vignettes about ophthalmology | Expert consensus | Low | Low | Low | Low | High |
| ^37^ | 2023 | Sorin | GPT-4V | Not written | Clinical vignettes about patients with ocular symptoms | Expert consensus | Low | Low | Low | Low | High |
| ^38^ | 2023 | Delsoz | GPT-3.5, GPT-4 | Not written | Clinical vignettes about ophthalmology | Expert consensus | Low | Low | Low | Low | High |
| ^39^ | 2023 | Levine | GPT-3 | Not written | Hypothetical clinical vignettes representing various common complaints | Expert consensus | Low | Low | Low | Low | High |
| ^40^ | 2023 | Schubert | GPT-4V | Not written | The quiz “Image Challenge” in NEJM | Answer | Low | High | Low | Low | High |
| ^41^ | 2023 | Sultan | GPT-3.5 | Not written | Clinical vignettes about patients with cancer predisposition syndromes | Expert consensus | Low | Low | Low | Low | High |
| ^42^ | 2023 | Kiyohara | PaLM2, GPT-3.5, GPT-4 | Not written | Clinical vignettes about vasospastic angina and acute coronary syndrome | Expert consensus | Low | Low | Low | Low | High |
| ^43^ | 2023 | Horiuchi | GPT-4 | August 3, 2023 | The quiz “Case of the Week” in AJNR | Answer | Low | High | Low | Low | Low |
| ^44^ | 2023 | Stoneham | GPT-4 | Not written | Clinical vignettes about patients with cutaneous symptoms | Expert consensus | Low | Low | Low | Low | High |
| ^45^ | 2023 | Rundle | GPT-3.5 | May 24, 2023 | Hypothetical clinical vignettes representing cutaneous tumors | Expert consensus | Low | Low | Low | Low | High |
| ^46^ | 2023 | Rojas-Carabali | GPT-3.5, GPT-4, Glass | Not written | Clinical vignettes about patients with uveitis | Expert consensus | Low | Low | Low | Low | High |
| ^47^ | 2023 | Fraser | GPT-3.5, GPT-4 | Not written | Clinical vignettes about emergency medicine | Expert consensus | Low | Low | Low | Low | High |
| ^48^ | 2023 | Krusche | GPT-4 | Not written | Clinical vignettes about rheumatology | Expert consensus | Low | Low | Low | Low | Low |
| ^49^ | 2023 | Galetta | GPT-4 | Not written | Clinical vignettes about neurology | Expert consensus | Low | Low | Low | Low | High |
| ^50^ | 2023 | Delsoz | GPT-3.5 | Not written | Clinical vignettes about ophthalmology | Expert consensus | Low | Low | Low | Low | High |
| ^51^ | 2023 | Hu | GPT-4 | Not written | Clinical vignettes about ophthalmology | Expert consensus | Low | Low | Low | Low | High |
| ^52^ | 2023 | Abi-Rafeh | GPT-3.5 | Not written | Hypothetical clinical vignettes about plastic surgery | Expert consensus | Low | Low | Low | Low | High |
| ^53^ | 2023 | Koga | PaLM2, GPT-3.5, GPT-4 | Not written | Clinical vignettes about neuropathology | Expert consensus | Low | Low | Low | Low | High |
| ^54^ | 2023 | Xv | GPT-3.5 | Not written | Clinical vignettes about urology | Expert consensus | Low | Low | Low | Low | Low |
| ^55^ | 2023 | Reese | GPT-4 | Not written | Clinical vignettes from case reports in NEJM | Expert consensus | Low | Low | Low | Low | High |
| ^56^ | 2023 | Han | GPT-3.5, GPT-4, GPT-4V, Llama 2,  Med-42 | Not written | The quiz “Image Challenge” in NEJM and the quiz "Image of the Month" in JAMA | Answer | Low | High | Low | Low | Unclear |
| ^57^ | 2023 | Senkaiahliyan | GPT-4V | Not written | Clinical vignettes about various medical images | Expert consensus | Low | Low | Low | Low | High |
| ^58^ | 2023 | Williams | GPT-3.5 | March 1, 2023 | Clinical vignettes about emergency medicine | Expert consensus | Low | Low | Low | Low | Low |
| ^59^ | 2023 | Tenner | GPT-3.5 | Not written | Clinical vignettes from case reports in NEJM | Expert consensus | Low | Low | Low | Low | High |
| ^60^ | 2023 | Mori | GPT-4 | Not written | Clinical vignettes about imaging findings of SAPHO syndrome | Expert consensus | Low | Low | Low | Low | Low |
| ^61^ | 2023 | Mykhalko | GPT-3.5 | Not written | The quiz “Case Challenges” from the Medscape website | Answer | Low | High | Low | Low | High |
| ^62^ | 2023 | Andrade-Castellanos | GPT-3.5 | Not written | The quiz “Test yourself” from the American College of Physicians website | Answer | Low | High | Low | Low | High |
| ^63^ | 2023 | Daher | GPT-3.5 | Not written | Clinical vignettes about patients with joint symptoms | Expert consensus | Low | Low | Low | Low | High |
| ^64^ | 2023 | Suthar | GPT-4 | July 20, 2023 | The quiz “Case of the Month” in AJNR | Answer | Low | High | Low | Low | Low |
| ^65^ | 2023 | Nakaura | Prometheus, GPT-3.5 | Not written, June 13, 2023 | Clinical vignettes about various imaging findings | Expert consensus | Low | Low | Low | Low | High |
| ^66^ | 2023 | Berg | GPT-3.5, GPT-4 | Not written | Clinical vignettes about emergency medicine | Expert consensus | Low | Low | Low | Low | High |
| ^67^ | 2023 | Gebrael | GPT-4 | Not written | Clinical vignettes about patients with prostate cancer | Expert consensus | Low | Low | Low | Low | High |
| ^68^ | 2023 | Ravipati | GPT-3.5 | Not written | Clinical vignettes about dermatology | Expert consensus | Low | Low | Low | Low | High |
